# Covarying Amplitude Modulation and Pulse Rate Enhances Pitch Discrimination in Cochlear Implant Users

**DOI:** 10.64898/2025.12.03.25341217

**Authors:** Hongxin Li, Huali Zhou, Limin Pang, Juanjuan Li, Chaogang Wei, Peina Wu, Qinglin Meng, Xianhai Zeng

**Affiliations:** School of Physics and Optoelectronics, South China University of Technology, Guangzhou, 510641, China; School of Electronics and Information Engineering, Heyuan Polytechnic, Heyuan, 517000, China; Department of Otolaryngology-Head and Neck Surgery, Guangdong Cardiovascular Institute, Guangdong Provincial People’s Hospital, Guangdong Academy of Medical Sciences, 519041, China; Shenzhen Longgang Otolaryngology Hospital and Shenzhen Institute of Otolaryngology, Shenzhen, 518172, China; Shenzhen Longgang Otolaryngology Hospital-South China University of Technology Joint Laboratory for Digital Hearing Healthcare, Shenzhen 518172, China

**Keywords:** Cochlear implant, pitch discrimination, temporal fine structure, consonant recognition

## Abstract

Pitch plays a fundamental role in prosody, lexical tone, and music perception, yet cochlear implant (CI) users exhibit limited temporal pitch sensitivity, particularly at higher pulse rates (e.g., 300 Hz). This study proposes a covarying pitch-encoding method, where the amplitude-modulation (AM) frequency and the pulse rate vary together in predetermined integer ratios to reinforce temporal periodicity cues. A single-channel psychophysical pitch discrimination experiment was conducted at the most apical electrode in 17 ears from 14 Cochlear™ CI users around reference frequencies of 50 and 300 Hz. The aims were to examine the effects of the integer ratio on pitch discrimination using the covarying method and to compare the performance of the covarying method with two conventional pitch encoding methods—pulse rate only and AM only. An additional consonant recognition task evaluated speech recognition ability. Results showed that at 50 Hz neither integer ratio nor pitch encoding method significantly affected pitch discrimination thresholds (≈30%). At 300 Hz, thresholds were overall higher than at 50 Hz, but the covarying method produced lower thresholds (41.8%) than the pulse rate (52.1%) and AM frequency methods (52.4%), and the covarying-pulse-rate difference was statistically significant. These results suggest that covarying stimulation can modestly enhance pitch discrimination at higher frequencies relative to conventional methods. Regression analyses revealed that temporal pitch discrimination in this psychophysical task at 50 Hz deteriorated with longer CI experience, whereas consonant recognition in a more ecologically relevant speech task improved, suggesting distinct neural adaptation mechanisms.

## 1. Introduction

Pitch plays a vital role in auditory perception, supporting prosody, lexical tone identification in tonal languages, speaker segregation in noise, and music perception. In cochlear implant (CI) users, pitch can be conveyed via *place cues* (with more apical electrodes eliciting lower pitch) or *temporal cues* (with higher pulse rates or modulation frequencies yielding higher pitch) (Tong et al., 1982; Zeng, 2002; Landsberger and Galvin, 2011). Unlike normal-hearing (NH) listeners, CI users receive place and temporal cues in a more independent manner.

In theory, CI users can access various combinations of place and temporal cues, potentially enhancing pitch perception. In practice, however, they exhibit poorer discrimination than NH listeners. NH listeners can detect F0 changes around 100 Hz as small as 1–2% for complex tone or periodic click sequences, whereas CI users often require 5–40% changes (Goldsworthy, 2015; Kaernbach and Bering, 2001). The limitation arises from both the loss of temporal fine structure in most CI coding strategies and intrinsic temporal constraints in electric hearing (Carlyon et al., 2025). Notably, pitch discrimination typically deteriorates at pulse rates exceeding ∼300 pulses per second (pps), a phenomenon often referred to as the *upper temporal limit* of pitch perception with electric hearing (Tong and Clark, 1985; Zeng, 2002; Macherey and Carlyon, 2014).

Pulse rate has been shown to be better than AM frequency for CI pitch discrimination (Baumann and Nobbe, 2004; Goldsworthy et al., 2022), highlighting the importance of pitch-encoding methods. Motivated by pattern-matching models in NH pitch perception and the independent processing of pulse and AM frequency in CI users, a covarying method is proposed, where pulse rate and AM frequency share a fixed integer ratio relationship on a single electrode. The hypothesis is that this method may enhance pitch discrimination by reinforcing temporal periodicity cues. Three temporal pitch-encoding methods used in our experiment are illustrated in Fig. 1. In the pulse rate method, unmodulated pulse trains are delivered at different rates *r*. In the AM method, the pulse rate *r* is fixed at 6000 pps while the amplitude is modulated at different frequencies *f_m_*. In the covarying method, pulse rate varies in proportion to the modulation frequency, here in the demonstrations using a fixed integer ratio *r*/*f_m_* = 4:1.

**Fig. 1.**
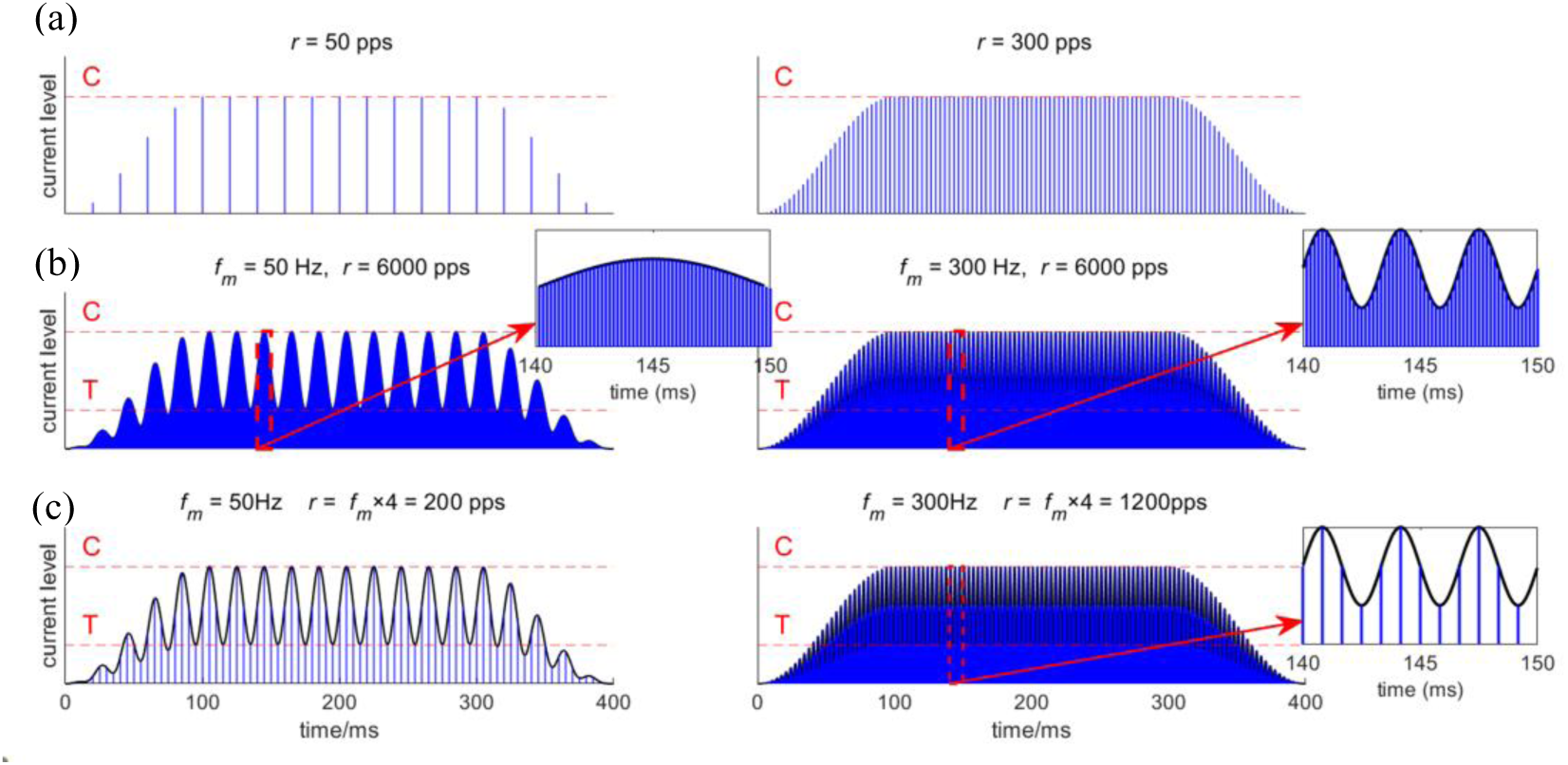
Illustrations of the three temporal pitch encoding methods. a) Pulse rate method (top): unmodulated pulse trains with different pulse rates. b) AM method (middle): the pulse rate *r* was fixed at 6000 pps and the amplitude was modulated at different modulation frequencies *f_m_*. c) Covarying method (bottom): the pulse rate varied proportionally with the modulation frequency (shown here with *r*/*f_m =_* 4:1).

Previous studies have demonstrated that modulation detection thresholds improve with increasing pulse rate (Galvin and Fu, 2005, 2009; Pfingst et al., 2007). Moreover, in the AM method, varying only the AM frequency may introduce stimulation distortion, which tends to diminish with higher pulse rates (Goldsworthy et al., 2021). Although the present study focuses on pitch discrimination rather than modulation detection, the two tasks are related in that both rely on accurate temporal coding of envelope cues. Therefore, a high stimulation rate of 6000 pps was adopted in the AM method to minimize potential distortion and ensure more robust encoding of temporal information. For the covarying method, different integer ratios of *r*/*f_m_* were tested, which in practice correspond to different stimulation rates for the same *f_m_*, allowing us to examine how pulse rate itself interacts with modulation frequency in shaping pitch discrimination performance.

Therefore, this study aims (1) to investigate the effect of the integer ratio of *r*/*f_m_* on pitch discrimination with the covarying method, and (2) to compare its performance with the conventional pulse-rate and AM-frequency encoding methods. In addition, a consonant recognition experiment was conducted as a measure of speech recognition ability, chosen in part because consonant identification relies heavily on temporal resolution and represents a more naturalistic, ecologically relevant listening condition.

## 2. Method

### 2.1 Participants

A total of 14 adult CI users using Cochlear^TM^ devices were recruited (17 ears), including 8 males and 6 females aged 23-47 years. The experiment lasted approximately 4 hours for each participant. All participants received monetary compensation and provided written informed consent prior to participation. This study was approved by the Institutional Review Board of Guangdong Provincial People’s Hospital. Participant details are provided in Table 1.

**Table 1.** Participant information.

| ID | Age range (Years) | Gender | Etiology | Tested ear | Implant Model | CI experience (Years) |
| --- | --- | --- | --- | --- | --- | --- |
| C1 | 26-30 | F | Ototoxicity | R | CI24R | 21-25 |
| C14 | 46-50 | F | Ototoxicity | R | CI24R | 15-20 |
| C24 | 26-30 | F | Ototoxicity | R | CI24M | 21-15 |
| C26 | 26-30 | M | Meningitis | R | CI24RE | 15-20 |
| C47 | 26-30 | F | Congenital | R | CI24R | 21-25 |
| C50 | 26-30 | F | Ototoxicity | L | CI24RE | 10-15 |
| C55 | 41-45 | M | Ototoxicity | L | CI512 | 1-5 |
| C56_L | 26-30 | F | Ototoxicity | L | CI512 | 1-5 |
| C56_R | 26-30 | F | Ototoxicity | R | CI512 | 1-5 |
| C57 | 26-30 | M | Congenital | L | CI622 | 1-5 |
| C58_L | 31-35 | F | Ototoxicity | L | CI512 | 1-5 |
| C58_R | 31-35 | F | Ototoxicity | R | CI512 | 1-5 |
| C60_L | 26-30 | M | Ototoxicity | L | CI512 | 6-10 |
| C60_R | 26-30 | M | Ototoxicity | R | CI24RE | 1-5 |
| C61 | 26-30 | M | Auditory neuropathy | L | CI622 | 1-5 |
| C62 | 26-30 | F | Ototoxicity | L | CI522 | 1-5 |
| C63 | 21-25 | M | EVAS | L | CI522 | 1-5 |

F: female; M: male; R: right ear; L: left ear. Participant IDs were assigned according to the internal database of our research group. As required by medRxiv, participant age and experience were presented as a range, but the actual years were used for analysis.

### 2.2 Stimuli

As demonstrated in Fig. 1, three pitch-encoding methods were investigated: unmodulated pulse trains (pulse rate method), amplitude-modulated pulse trains (AM method), and stimuli in which the AM frequency and pulse rate were kept at a fixed ratio (covarying method). For all modulated stimuli, the first pulse train starts at 0 phase of the sinusoidal modulator. Each stimulus lasted 400 ms, including 100 ms onset and offset ramps shaped as quarter-cycle sine waves. All stimuli consisted of biphasic current pulses with a phase duration of 25 µs and an interphase gap of 8 µs, and were delivered to electrode 22 (the most apical electrode) in MP1+2 mode via the Nucleus Implant Communicator (NIC4) (Gransier et al., 2024; Litovsky et al., 2017) connected to a personal computer running custom MATLAB software.

### 2.3 Procedure

#### 2.3.1 Loudness balance

To provide the basis for subsequent stimulus-level control, threshold (T) and most comfortable (C) levels were first measured across a broad range of pulse rates (40, 50, 100, 200, 300, 400, 500, 600, 1200, 2400, 3000, and 6000 pps). These measurements were later used to optimize stimulus intensity selection and to minimize potential loudness cues in the pitch discrimination tasks. Threshold estimation was carried out using an adaptive bracketing procedure. Current levels were first adjusted to ensure audibility, then decreased in 2 current unit (CU) steps until the stimulus was no longer perceived, followed by increases in 1-CU steps until the stimulus was reported as audible. The decrease–increase cycle was repeated, and each time the level at which audibility was regained was recorded. The T level was defined as the first current level that was reported as audible on two separate ascending sequences. During the test, loudness was categorized into four levels: *very soft*, *medium*, *loud*, and *too loud and uncomfortable*. On each presentation, the listener was required to indicate the perceived loudness category. The T level was first measured, after which the stimulation current was gradually increased using step sizes that decreased progressively, until the first time the listener reported *too loud and uncomfortable*; the level was recorded as the C level. For untested intermediate pulse rates between 40 and 6000 pps, the T and C levels were estimated by linear interpolation on the logarithmic scale of pulse rates.

#### 2.3.2 Pitch discrimination

Pitch discrimination thresholds were measured around reference frequencies or rates of 50 and 300 Hz (or pps), with the reference values uniformly jittered within a range of ± one-quarter octave to prevent listeners from memorizing a fixed reference frequency. The experiment consisted of three repeated-measures blocks, each containing eight conditions presented in a randomized order. Thresholds obtained from the three repetitions were averaged for analysis. At 50 Hz, five conditions were tested: 1) the pulse rate method varying pulse rate *r* with unmodulated amplitude; 2) the AM method varying the modulation frequency *f_m_* with a fixed *r* = 6000 pps; 3–5) the covarying method varying *f_m_* while fixing the *r*/*f_m_* ratios at 4, 10, or 30 respectively. At 300 Hz, three conditions were tested: 1) the pulse rate method varying pulse rate *r* with unmodulated amplitude; 2) the AM method varying the modulation frequency *f_m_* with a fixed *r* = 6000 pps; 3) the covarying method varying both *f_m_* and *r* while fixing the *r*/*f_m_* ratios at 4. For all modulated stimuli, modulation depth was set to span the dynamic range, with the peak current level corresponding to the C level and the theoretical trough level corresponding to the T level. For unmodulated stimuli, the current level was set to the C values.

A two-interval, two-alternative forced-choice (2I-2AFC) paradigm was used. Each trial consisted of a reference and a target stimulus presented in random order, separated by a 200-ms interstimulus interval. The listener was asked to indicate which interval contained the stimulus with the higher pitch. The target stimulus always had a higher modulation frequency *f_m_* (or pulse rate *r*) than the reference, with the assumption that the stimulus with the higher *f_m_* or *r* corresponds to a higher perceived pitch. Feedback was provided after each response. To minimize loudness cues, the peak current level of both the reference and target stimuli was randomly roved between 90% and 100% of the dynamic range above the T level defined for their respective pulse rates. Each waveform was linearly mapped within the individual dynamic range so that its peak corresponded to the roved level while preserving the original envelope shape. The roving was applied before the onset and offset ramps were imposed.

The initial parameter difference in *r* (for the pulse rate method) or *f_m_* (for the other two methods) between the reference and target stimuli was set to 100%. An adaptive procedure with weighted step sizes was used to target the 75% correct threshold (Kaernbach, 1991). If the response was correct, the parameter difference was reduced; if incorrect, the difference was increased. If the difference exceeded 100%, it was capped at 100%. Before the fourth reversal, correct responses decreased the difference by a factor of 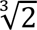, while incorrect responses increased it by a factor of 2. After that, before the eighth reversal, correct responses decreased the difference by 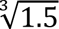, and incorrect responses increased it by 1.5. For all subsequent reversals, the step sizes were 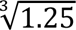 (down) and 1.25 (up). A run terminated after eight incorrect responses and also terminated early if the participant made five incorrect responses with a difference of 100%. The discrimination threshold was defined as the mean percentage parameter difference at the last four reversal points relative to the reference.

### 2.4 Consonant recognition

Consonant recognition was tested using a monosyllabic consonant corpus comprising 89 /CV/ syllables to provide an additional measure of speech recognition ability. The stimuli were organized into minimal contrast sets containing three or four items that shared the same vowel but differed in the initial consonant, covering major Mandarin consonant categories including plosives, affricates, fricatives, nasals, and approximants. Consonants were combined with five vowel contexts (/a/, /i/, /u/, /e/, /o/) to form sets such as 渣zhā, 插chā, 杀shā, rā (/a/ context) and 机 jī, 七 qī, 西 xī (/i/ context). The corpus included both meaningful and nonsense syllables (Supplementary Table S1) and all stimuli were produced by a female native speaker. Consonant identification was chosen in part because it is largely influenced by temporal resolution, particularly for fricatives and affricates that contain rapid envelope fluctuations, thereby serving as a more ecologically relevant behavioral measure complementary to the basic psychophysical pitch discrimination task.

Participants wore a CCI-Mobile device (Ghosh et al., 2022) working in the real-time mode using participants’ clinical MAPs. This platform was chosen because using a standardized research platform ensures cross-participant consistency by minimizing hardware and software differences across individual clinical devices. Testing took place in a sound-attenuated booth, with a loudspeaker positioned 1 m directly in front of the participant. Prior to testing, the presentation level was individually adjusted to a comfortable level. The test was administered via a graphical user interface developed in MATLAB. On each trial, listeners selected the perceived consonant from either three or four alternative choices, yielding a total of 89 trials. No feedback on response accuracy was provided.

## 3. Result

The three repeated blocks of pitch discrimination thresholds showed no significant differences (Friedman statistic = 3.25, *p* = 0.236), supporting the use of the mean value across blocks for further analyses.

### 3.1 Dynamic range

Measured T and C levels from 17 ears were averaged and plotted as functions of pulse rate in Fig. 2. Both T and C levels decreased with increasing pulse rate. However, the dynamic range became slightly narrower from 40 to 300 pps and wider from 300 to 6000 pps. Table 2 shows the average dynamic range at 17 different pulse rates from 40 to 6000 pps.

**Fig. 2.**
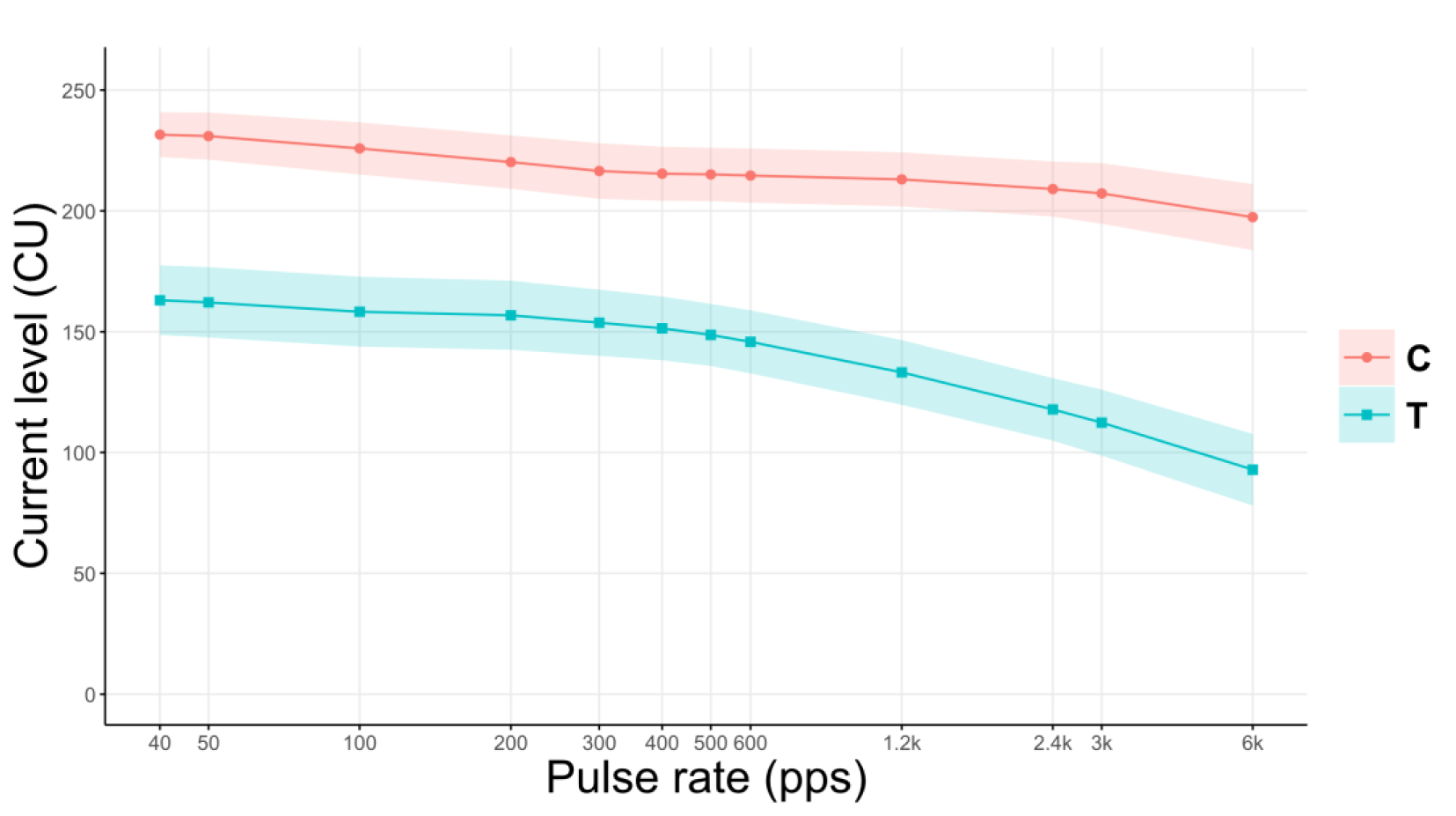
Mean T (lower curve) and C (upper curve) levels across pulse rates for all 17 ears at the most apical electrode. Shaded areas represent 95% confidence intervals.

**Table 2.** Mean threshold (T) and most comfortable (C) levels, and corresponding dynamic ranges (mean ± SD) across pulse rates.

| Pulse rate<br>(pps) | T level<br>(CU) | C level<br>(CU) | Dynamic range<br>(CU) | SD<br>(CU) |
| --- | --- | --- | --- | --- |
| 40 | 163 | 232 | 69 | 22 |
| 50 | 162 | 231 | 69 | 22 |
| 100 | 158 | 226 | 68 | 20 |
| 200 | 157 | 220 | 63 | 20 |
| 300 | 154 | 217 | 63 | 20 |
| 400 | 151 | 215 | 64 | 18 |
| 500 | 149 | 215 | 66 | 19 |
| 600 | 146 | 215 | 69 | 19 |
| 1200 | 133 | 213 | 80 | 20 |
| 2400 | 118 | 209 | 91 | 19 |
| 3000 | 112 | 207 | 95 | 21 |
| 6000 | 93 | 197 | 104 | 22 |

### 3.2 Effect of the integer ratio in the covarying method on pitch

For the covarying method at a reference frequency of 50 Hz, the mean pitch discrimination thresholds were 29.3%, 27.3%, and 26.9%, for *r*/*f_m_* ratios of 4, 10, and 30, respectively. The data under the three ratio conditions all passed the Shapiro–Wilk normality test, with *p*-values of 0.0820, 0.2437, and 0.2533 respectively. Figure 3 shows the average and individual thresholds across these three ratios. Greenhouse–Geisser correction (*ε* = 0.7594) was applied to adjust for violations of sphericity. A one-way repeated-measures ANOVA was conducted on the discrimination thresholds with *r*/*f_m_* ratio as a factor, revealing no significant effect (*F*_1.519, 24.30_ = 0.6172, *p* = 0.5048). Substantial inter-subject variability was observed, with individual thresholds ranging from as low as 5.3% to as high as 59.0%.

**Fig. 3.**
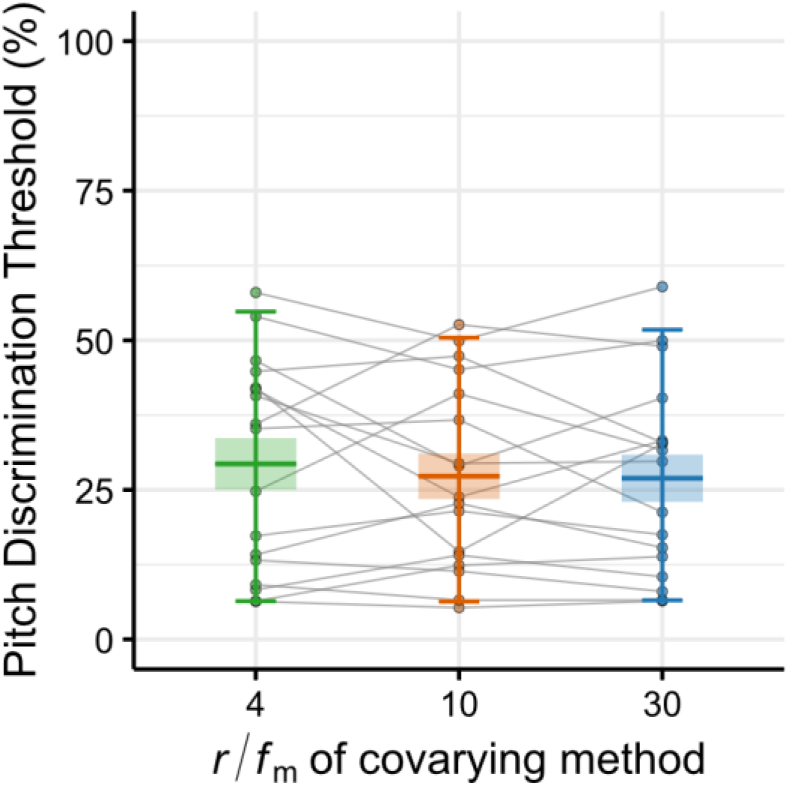
Pitch discrimination thresholds for the three *r*/*f_m_* ratios (4, 10, 30) at 50 Hz. Individual data points are shown as dots. Data points from the same participant are connected by solid lines. The central lines in the box plot represent the mean, and the boxes represent the standard errors of the means. The whiskers extend to the 5th and 95th percentiles of the data, encompassing the central 90% of the distribution.

### 3.3 Effect of pitch encoding method on pitch discrimination threshold

As no significant effect of the integer ratio was found at 50 Hz (Section 3.2), the thresholds from the three ratios (4, 10, 30) were averaged to represent the covarying method at this frequency. At 300 Hz, only the 4:1 ratio was tested; thus, no averaging was necessary. These covarying results were then compared with those from the pulse-rate and AM-frequency methods. Figure 4 shows the results with three methods at 50 Hz and 300 Hz. At 50 Hz, mean thresholds were 27.9%, 29.1%, and 28.9% for the covarying, pulse rate, and AM frequency methods, respectively. At 300 Hz, the corresponding values were 41.8%, 52.1%, and 52.4%.

**Fig. 4.**
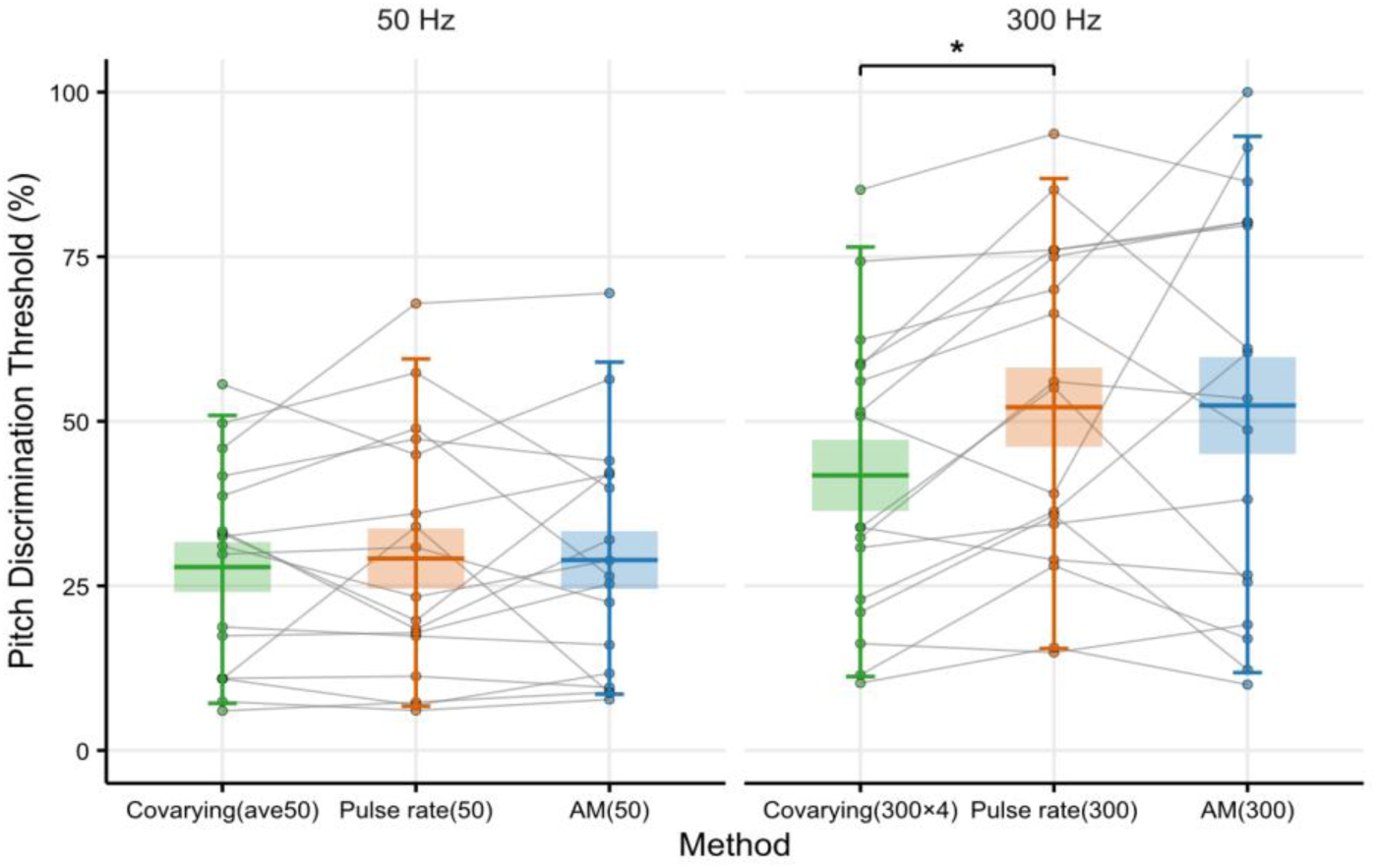
Pitch discrimination thresholds of the three encoding methods at reference frequency of 50 Hz (left) and 300 Hz (right). Individual data points are shown as dots. Data points from the same participant are connected by solid lines. The central lines in the box plot represent the mean, and the boxes represent the standard errors of the means. Asterisks (*) indicate a statistically significant difference (*p* < 0.05). The whiskers extend to the 5th and 95th percentiles of the data, encompassing the central 90% of the distribution.

Data from all six conditions shown in Fig. 4 passed the Shapiro–Wilk test for normality, with *p*-values of 0.2813, 0.2402, 0.1821, 0.4784, 0.2972, and 0.1672 from left to right. A two-way repeated-measures ANOVA with interactions was conducted on the discrimination thresholds with pitch-encoding method and reference frequency as within-participant factors, followed by Tukey post-hoc multiple comparisons. Greenhouse–Geisser correction (*ε* =0.7798) was applied to adjust for violations of sphericity. Results showed a significant difference between 50 Hz and 300 Hz (*F*_1, 32_ = 8.025, *p* = 0.0079), among pitch encoding methods (*F*_1.560, 49.90_ = 3.929, *p* = 0.0354) but no significant effect of the interaction between pitch-encoding method and reference frequency (*F*_2, 64_ = 2.505, *p* = 0.0897). Specifically, at 50 Hz, there was no significant effect of encoding methods (covarying versus pulse rate, *p* = 0.8704; covarying versus AM, *p* = 0.8588; pulse rate versus AM, *p* = 0.9969). Substantial inter-subject variability was observed, with individual thresholds ranging from as low as 6.0% to as high as 69.5%. At 300 Hz, the covarying method yielded a mean threshold that was 10.6 percentage points lower than that in the AM method, although the difference was not significant (*p* = 0.0525), and 10.3 percentage points lower than that in the pulse rate method, which was significant (*p* = 0.0031). Mean thresholds in the pulse rate method were only 0.2 percentage points lower than those in the AM method, and the difference did not reach significance (*p* = 0.9986). Although the covarying–AM difference was not statistically significant, 12 of the 17 ears exhibited lower thresholds with the covarying method than with the AM method. Overall, thresholds were generally lower with the covarying method compared to the pulse rate and AM methods, reflecting the trend seen in the majority of participants.

### 3.4 Threshold as a function of CI experience

Individual differences in pitch discrimination performance may be influenced by listener-related factors such as CI experience or age. A linear bivariate regression model of the form *y* = *β*_0_ + *β*_1_*x*_1_ + *β*_2_*x*_2_ + *∈* was first employed to evaluate the influence of CI experience and age on pitch discrimination thresholds (*n* = 17). In the model, *x*_1_ and *x*_2_ represent CI experience (years) and age, respectively; *β* denotes the regression coefficients, and *∈* represents the residual error. As shown in the bivariate results (Table 3), at 50 Hz, the model showed significant negative effects of CI experience on pitch discrimination for all three methods (*p* < 0.05). Age also had a significant negative effect on pitch discrimination for the covarying and pulse rate methods (*p* < 0.05), but no significant effect for the AM method (*p* = 0.4820). At 300 Hz, no statistically significant effects were found for most methods, though a positive correlation trend was observed.

**Table 3.** Results of linear regression analyses on pitch discrimination thresholds as a function of CI experience and age.

| Condition | Ears<br>(*n*) | Bivariate<br>(CI experience and age) |  |  | Univariate<br>(CI experience) |  |
| --- | --- | --- | --- | --- | --- | --- |
| | | $R^2$ | $p$<br>(CI experience) | $p$<br>(age) | $R^2$ | $p$<br>(CI experience) |
| Covarying<br>(ave50) | 17 | 0.638 | **0.0006\*** | **0.0221\*** | 0.467 | **0.0025\*** |
|  | 11 | 0.711 | 0.0662 | 0.2600 | 0.659 | **0.0024\*** |
| Pulse rate<br>(50) | 17 | 0.656 | **0.0003\*** | **0.0288\*** | 0.510 | **0.0013\*** |
|  | 11 | 0.815 | **0.0026\*** | 0.8220 | 0.813 | **0.0001\*** |
| AM (50) | 17 | 0.531 | **0.0015\*** | 0.4820 | 0.513 | **0.0012\*** |
|  | 11 | 0.685 | **0.0363\*** | 0.6030 | 0.674 | **0.0020\*** |
| Covarying<br>(300×4) | 17 | 0.203 | 0.3510 | 0.1150 | 0.043 | 0.4260 |
|  | 11 | 0.399 | 0.5720 | 0.2880 | 0.302 | 0.0801 |
| Pulse rate<br>(300) | 17 | 0.083 | 0.4680 | 0.3910 | 0.032 | 0.4950 |
|  | 11 | 0.355 | 0.5750 | 0.3500 | 0.275 | 0.0975 |
| AM (300) | 17 | 0.426 | 0.5080 | **0.0066\*** | 0.010 | 0.7040 |
|  | 11 | 0.391 | 0.6380 | 0.0942 | 0.117 | 0.3040 |

Significance levels: \* $p < 0.05$ .

Among the 17 ears, 11 were aged 27–30 years, with a wide range of CI experience (3 – 25 years). By restricting the bivariate analysis to this age-homogeneous subgroup (*n* = 11), the potential confounding effect of age was largely minimized, and a strong negative effect of CI experience on pitch discrimination for pulse rate and AM methods was still observed at 50 Hz.

For clarity and ease of visualization, the results of the bivariate regression model were not plotted. Instead, a univariate linear regression model was used, with CI experience as the predictor, to illustrate its relationship with pitch discrimination thresholds (Figure 5). This approach was applied to both the full sample of 17 ears and the age-homogeneous subgroup of 11 ears. The corresponding statistical results are provided in the univariate section of Table 3.

**Fig. 5.**
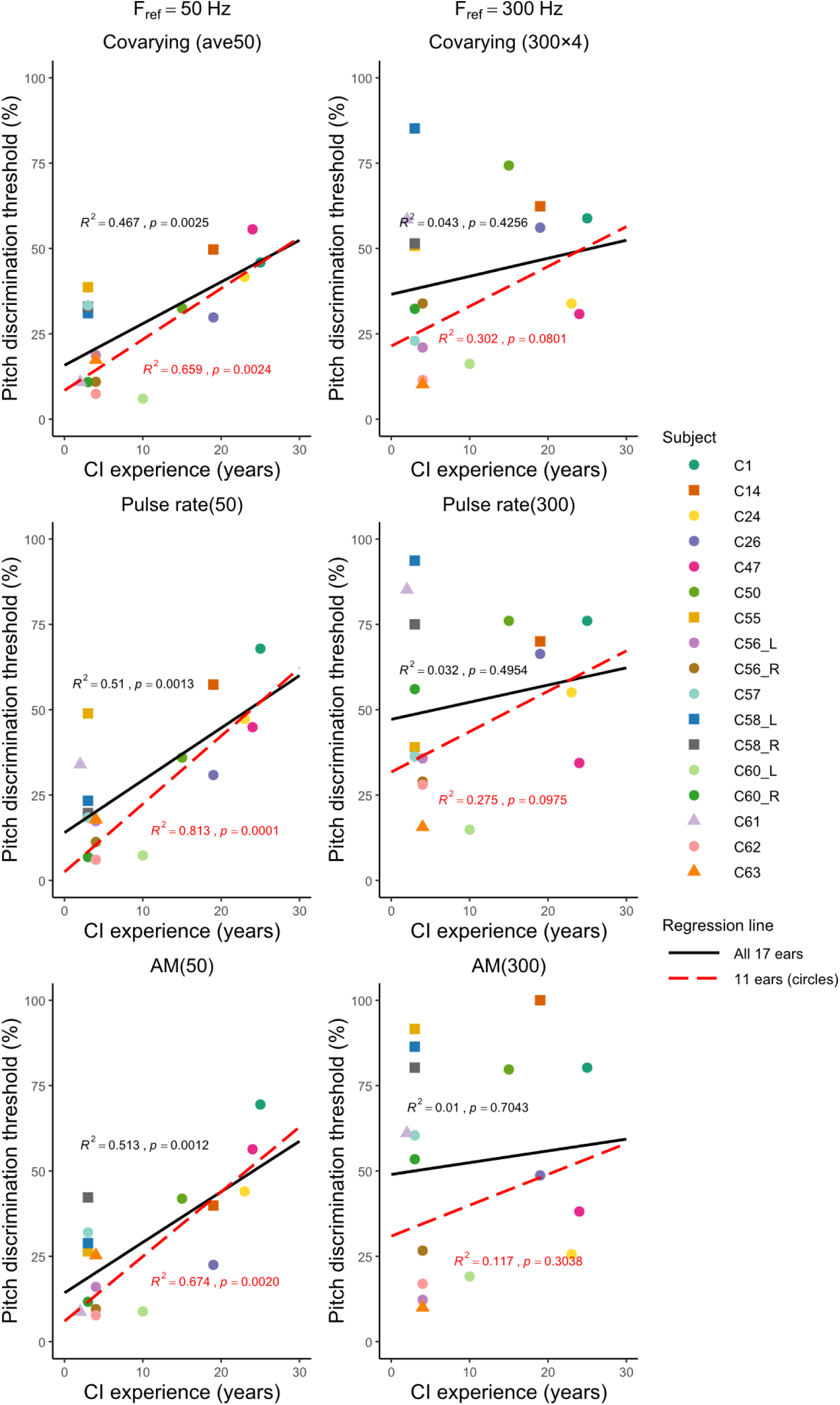
Pitch discrimination thresholds as a function of CI experience for the covarying, pulse rate, and AM methods. The solid black line represents the regression fit for all 17 ears, and the red dashed line represents the fit for the subgroup of 11 ears aged 27–30 years. Circular markers denote these 11 ears, and square markers denote the four older ears, triangle markers indicate ears younger than 27 years. The coefficient of determination (*R*²) and significance (*p*) are shown for the fit.

Figure 5 illustrates pitch discrimination thresholds as a function of CI experience across the three encoding methods. The results were consistent with those from the above bivariate regression model, showing significant positive correlations with pitch discrimination threshold at 50 Hz and nonsignificant but positive trends at 300 Hz. These findings suggest that temporal pitch discrimination ability may deteriorate with increasing CI experience.

Moreover, by averaging the slopes of the three regression lines at 50 Hz, the rate constant was estimated to be *β*_1_ ≈ 1.4, suggesting that pitch discrimination threshold increases at an average rate of approximately 1.4% per year. This indicates that the change evolves at a very slow rate over time. However, because the present data are cross-sectional rather than longitudinal, this rate should be interpreted as a correlational trend rather than a true temporal progression.

Figure 6 shows consonant correct rate as a function of CI experience. Following the same approach, data from all 17 ears were first fitted with the bivariate regression model, which showed no significant effect of age (*p* = 0.8132), and were then analyzed using the univariate regression model. Consonant recognition exhibited a significant positive correlation with CI experience (17 ears: *R*² = 0.297, *p* = 0.0238; 11 ears: *R*² = 0.366, *p* = 0.0486). Because consonant identification reflects a more naturalistic and ecologically relevant speech task than basic psychophysical measures, this finding highlights functional improvements associated with long-term auditory training in daily communication contexts.

**Fig. 6.**
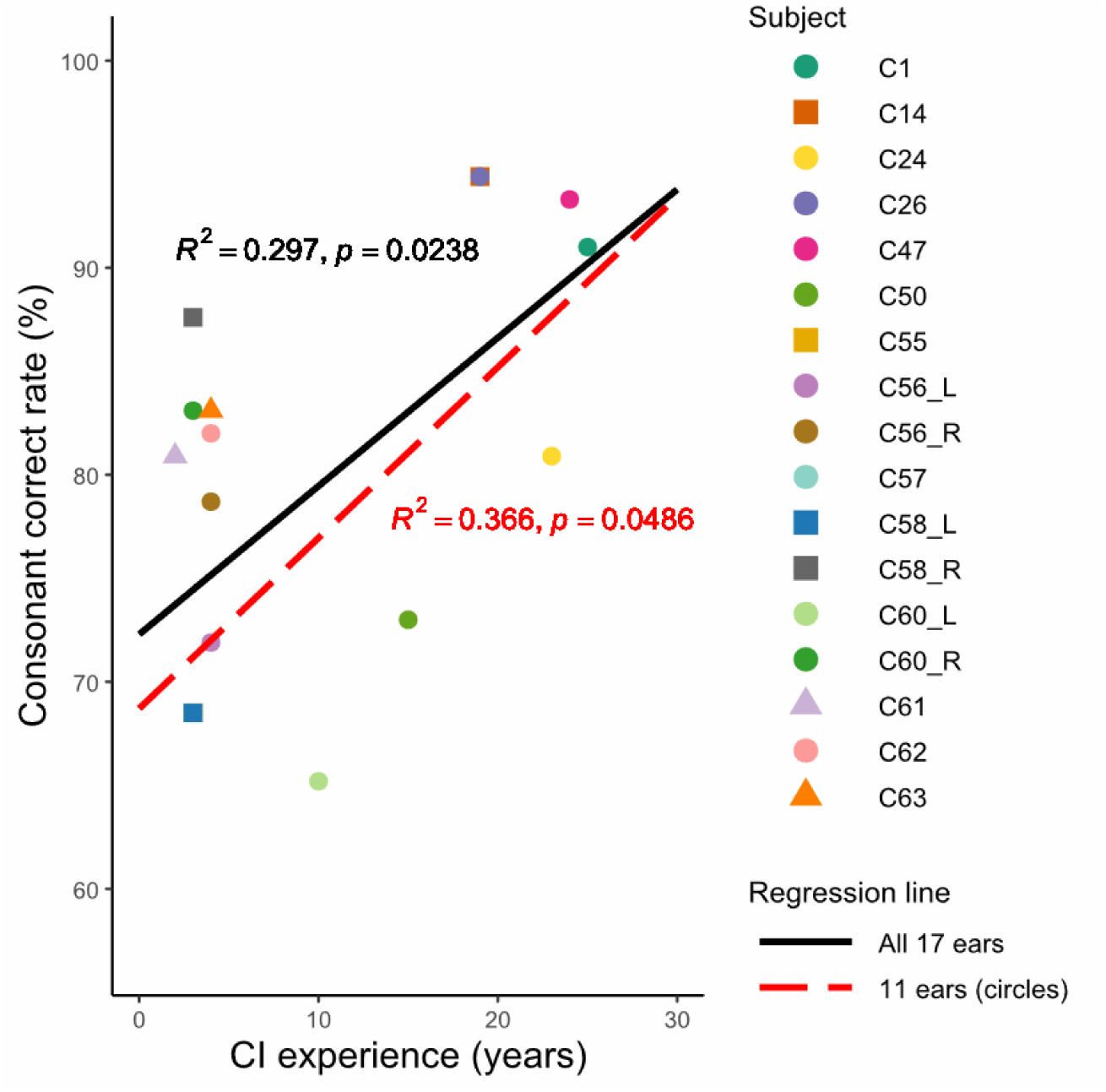
Consonant correct rate as a function of CI experience. Circular markers indicate ears aged 27–30 years (*n* = 11), triangle markers indicate ears younger than 27 years (*n* = 2), and square markers indicate ears older than 30 years (*n* = 4). The coefficient of determination (*R*²) and significance (*p*) are shown for the fit.

## 4. Discussion

Given the crucial role of pitch in auditory perception and the generally poor pitch performance observed in CI users, a covarying temporal pitch coding method was proposed to improve pitch discrimination. Through the experiments reported in this study, two questions have been addressed: (1) how the integer ratio affects the performance of the covarying pitch coding method, and (2) how this temporal pitch coding method compares with the two conventional methods, AM and pulse rate, in terms of providing pitch discrimination ability. Furthermore, we examined how temporal pitch resolution relates to CI experience, and found a somewhat unexpected negative association between CI experience and temporal pitch discrimination ability.

### 4.1 Comparison of pitch discrimination between pulse rate and AM methods

In the present study, no significant difference in pitch discrimination was observed between the pulse rate and AM methods, consistent with the findings of Kong et al. (2009). In contrast, Baumann and Nobbe (2004) and Goldsworthy et al. (2022) demonstrated pitch discrimination thresholds with the pulse rate method were significantly better than with the AM method. Goldsworthy et al. (2022) tested eight participants (12 ears) and found that pulse rate yielded significantly better pitch discrimination at 220 Hz and 440 Hz (but not 110 Hz), with 9 out of 12 ears favoring pulse rate over AM frequency. Kong et al. (2009) tested five participants over a range of 100–500 Hz, where only one participant consistently performed better in the pulse rate method; others showed no clear pattern, possibly due to methodological differences. Baumann and Nobbe (2004) tested only three participants in the range of 200–566 Hz, where significant results may be affected by sampling bias. The discrepancies with the present study may be attributable to differences in participant number, frequency points tested, experimental procedures, loudness balancing methods, or individual interpretations of T and C levels, which may affect perceived loudness of modulated stimuli.

Notably, pitch discrimination thresholds in the present experiment were higher than those reported by Goldsworthy et al. (2022). At 300 Hz, the present thresholds of 52.1% (pulse rate) and 52.4% (AM), compared with 33.3% (pulse rate) and 51.1% (AM) at 440 Hz in their study, potentially due to training effects (Goldsworthy and Shannon, 2014). However, the range of thresholds observed in the present experiment was broadly in agreement with other studies. At 50 Hz, the present study yielded mean thresholds of 29.1%, compared with the 24% pulse rate threshold reported by Zeng et al. (2002). At 300 Hz, the present threshold (52.4%) is comparable to the AM threshold of >56.5% at 283 Hz reported by Baumann and Nobbe (2004).

### 4.2 Enhanced pitch discrimination with the covarying method

At 50 Hz, the experimental results showed no significant differences among the three pitch encoding methods. Previous studies have shown that, at a fixed modulation frequency of 100 Hz, the effect of different pulse rates on pitch perception is non-monotonic (McKay et al., 1994), meaning that increases in pulse rate do not necessarily produce higher perceived pitch. Such non-monotonicity may lead to fluctuations in perceived pitch under the covarying method, thereby limiting pitch discrimination performance. Moreover, factors such as the choice of reference frequency (Goldsworthy et al., 2022) and electrode selection (Baumann and Nobbe, 2004) may also have contributed to the lack of significant effects.

At 300 Hz, a significant difference in pitch-discrimination thresholds was observed only between the covarying and pulse rate methods. Thresholds were also higher than at 50 Hz, indicating reduced pitch sensitivity at higher reference frequencies. Consequently, pitch discrimination at 300 Hz appears more likely amenable to improvement. In contrast, pitch discrimination at 50 Hz was already quite good, with some participants achieving thresholds as low as 6.0%, leaving relatively limited room for further enhancement. Moreover, because the present dataset included only 17 ears, the sample size may have been insufficient to reveal potential differences among the methods at 50 Hz.

### 4.3 CI experience with consonant recognition and pitch discrimination

Although many auditory perceptual abilities improve with longer CI use due to adaptive plasticity—as reflected in our consonant recognition results—temporal pitch discrimination showed the opposite trend, with longer implantation associated with poorer performance. Zhou et al. (2019) has examined the relationship between pulse rate discrimination at 200, 300, and 500 pps and duration of implant use, but no clear trend was observed, indicating that pulse rate discrimination ability could not be reliably predicted from the duration of implantation. However, this study involved relatively older participants (57–77 years) with a limited range of implant durations (2.7–13.8 years). Gfeller et al. (2007) reported a statistically significant correlation between pitch ranking performance and months of implant use (*n* = 114, *r* = –0.25), but the correlation was weak given the small magnitude of *r*.

Several mechanisms may account for this decline. One possible interpretation is that, over extended periods, exposure to inaccurate temporal fine-structure cues in clinical coding strategies may drive maladaptive plasticity. Schnupp and colleagues have proposed that the weak sensitivity to ITDs in CI users may, in part, be due to the brain “losing interest” in temporal fine-structure information after prolonged exposure to unreliable or distorted cues (Buck et al., 2023; Carlyon et al., 2025). Another possible explanation lies in the nature of the experimental task. Gfeller et al. (2007) employed a relatively complex pitch-ranking paradigm, whereas the present study used a more basic single-electrode psychophysical task, which may provide a more direct measure of temporal pitch sensitivity. Moreover, perfect charge balance cannot be achieved with biphasic electrical pulses, and prolonged exposure to imperfectly balanced stimulation can lead to cochlear tissue changes and neural damage (Shepherd et al., 1993, 1991). This finding also indicates a decline in neural sensitivity with longer implant experience.

### 4.4 Limitations

Several limitations of the present study should be acknowledged. First, whether pitch conveyed by the same periodicity in pulse rate and AM frequency is perceived as identical remains a matter of debate. When pulse rate and AM frequency are not in an integer relationship, pitch percepts are generally similar; however, when such a relationship exists, AM stimuli may produce slightly higher pitch percepts than pulse rate stimuli (McKay et al., 1994). This potential discrepancy was not directly examined in the current study. Second, modulation detection thresholds should ideally be assessed before pitch discrimination with modulated pulse trains, but due to time constraints, this was not feasible, and all AM stimuli were presented at 100% modulation depth. Previous work has demonstrated that pitch discrimination improves with increasing modulation depth (Camarena and Goldsworthy, 2024). Third, in this study, modulated stimuli were presented between each participant’s T and C levels, whereas unmodulated stimuli were set at the mean current of T and C. This approach does not ensure equal loudness between modulated and unmodulated stimuli, nor across different methods (Fraser and McKay, 2012). Nonetheless, level roving was employed to minimize loudness cues, and participants consistently reported the stimuli as being of moderate loudness. It remains impossible to fully exclude the influence of loudness on pitch discrimination.

The present findings are limited to psychophysical measurements with single-electrode stimulation, and their clinical applicability remains uncertain. Electrode-specific factors such as insertion depth (Berg et al., 2025; Finley et al., 2008), frequency–place mapping (DeFreese et al., 2025), and neural health (Jeng et al., 2009; Shader et al., 2020) may alter the outcomes, so results obtained under single-electrode conditions may not directly generalize to multi-electrode stimulation. In multi-electrode contexts, channel interaction, spectral blurring (Tang et al., 2011), electrode activation order (Todd and Landsberger, 2018), inter-electrode asynchrony (De Groote et al., 2025), and virtual-channel effects (Donaldson et al., 2005) further complicate pitch encoding. Moreover, implementing covarying modulation rate and stimulation rate in real-time clinical coding strategies remains highly challenging. This highlights a gap between controlled laboratory manipulations and practical sound coding schemes in clinical CI systems.

## 5. Conclusion

The covarying encoding method proposed in this study, which maintains a fixed integer relationship between amplitude modulation frequency and pulse rate, has the potential to improve temporal pitch discrimination in CI users. Future work will continue to explore this covarying method, with a focus on advancing simultaneous multi-electrode stimulation and refining encoding methods. Although not a primary research aim, the observed decline of temporal pitch discrimination with CI experience provides secondary insights into the long-term perceptual consequences of current CI coding strategies.

## Data Availability

All data produced in the present study are available upon reasonable request to the authors

## Acknowledgments

We thank the Joint Laboratory of Digital Hearing Healthcare between Shenzhen Longgang Otolaryngology Hospital and South China University of Technology. Special thanks are extended to Cochlear Ltd. for their generous equipment support.

## CRediT authorship contribution statement

**Hongxin Li: Writing – original draft,** Writing – review and editing, Data curation, Formal analysis, Validation, Visualization, Conceptualization, Software. **Huali Zhou:** Formal analysis, Methodology, Writing – review and editing. **Limin Pang:** Data curation, Investigation, Resources. **Juanjuan Li:** Resources, Writing – review and editing, Supervision, Funding acquisition. **Chaogang Wei:** Supervision, Writing – review and editing, Data curation, Visualization. **Peina Wu:** Supervision, Resources, Funding acquisition. **Qinglin Meng:** Conceptualization, Formal analysis, Funding acquisition, Resources, Supervision, Writing – original draft, Writing – review and editing. **Xianhai Zeng:** Resources, Writing – review and editing, Supervision, Funding acquisition.

## Author Declarations

The authors have no conflicts to disclose.

## Data Availability

The datasets generated during and/or analyzed during the current study are available from the corresponding author on reasonable request.

## Declaration of generative AI and AI-assisted technologies in the manuscript preparation process

During the preparation of this work the author(s) used ChatGPT (GPT-5, OpenAI) in order to enhance the language and readability of the manuscript. After using this tool, the author(s) reviewed and edited the content as needed and take(s) full responsibility for the content of the published article.

